# Long-read sequencing enables high-accuracy mitochondrial heteroplasmy detection in Parkinson’s disease

**DOI:** 10.64898/2026.06.11.26355258

**Authors:** Theresa Lüth, Susen Schaake, Christoph Much, Mary Megan Belyea, Philip Seibler, Anne Grünewald, Patrick May, Christine Klein, Hansi Weissensteiner, Joanne Trinh

## Abstract

**Background:** Low-frequency heteroplasmic mitochondrial DNA (mtDNA) variants are associated with aging and neurological diseases, including Parkinson’s disease (PD). Targeted deep mtDNA sequencing using PacBio HiFi long reads has the potential to resolve heteroplasmy across the full mitochondrial genome with high accuracy.

**Methods:** To validate Vega PacBio sequencing for detecting mtDNA heteroplasmy, we analyzed four predefined mixtures of two mtDNA haplotypes. We generated a single long-range PCR amplicon covering the entire mitochondrial genome. These amplicons were mixed at predefined ratios (minor mixture haplotype component: 5%, 2%, 1%, and 0.1%). Variant calling was performed using *Mutserve2*, and accuracy was assessed by calculating the F_1_ score from comparisons between expected and detected variants. Full-length mtDNA PacBio sequencing was applied to investigate heteroplasmy across fibroblast passages derived from five LRRK2 p.Gly2019Ser variant carriers (n=3 affected with PD and n=2 unaffected carriers). Changes in mtDNA heteroplasmy level and variant load were assessed longitudinally using a linear mixed model.

**Results:** The single-amplicon approach enabled full-length haplotype resolution without amplification bias associated with overlapping PCR strategies. The F_1_ score of the predefined mixtures was 1.0 for heteroplasmy levels between 5% and 1% and remained high (0.91) at 0.1%. We detected n=10/62 variants discordant with the Illumina reference at the 0.1% mixture, but sensitivity remained very high at 1.00 in that mixture. Detected minor variants closely matched expected heteroplasmy levels, with average variant levels of 0.057 (5%), 0.022 (2%), 0.011 (1%), and 0.001 (0.1%). Across twelve fibroblast passages, we observed fewer mtDNA heteroplasmic variants (β=-3.2, p=0.026). Increased heteroplasmic variant load over time was also associated with older age (β=1.50, p=0.001) and PD affection status (β=5.0, p=1.0 × 10^-^⁴) in *LRRK2* variant carriers. Notably, we observed distinct patterns of heteroplasmic variants that either increased or decreased in heteroplasmy level across passages.

**Conclusion:** PacBio HiFi sequencing, combined with a single-amplicon strategy, enables accurate full-length mtDNA heteroplasmy detection and longitudinal analysis, providing a valuable tool for studying mitochondrial variation and dynamics in disease.

## Background

Ongoing mutation and selection of mitochondrial DNA (mtDNA) in both the germline and soma generate mtDNA heteroplasmy - the coexistence of multiple different mitochondrial genotypes within a cell, tissue, or organism. Damage repair in mtDNA is less efficient than in nuclear DNA, resulting in an ∼10× higher mutation rate. Consequently, not all mtDNA copies share the same variants; those present in only a subset are called heteroplasmic variants(1, 2). It has been reported that mtDNA heteroplasmy is not only closely linked to aging(3), but also implicated in a range of diseases, including neurodegenerative disorders such as Parkinson’s disease (PD)(4–6). Therefore, accurate characterization and quantification of mtDNA heteroplasmic variants are essential for understanding mitochondrial biology and the role of mtDNA heteroplasmy in disease.

Next-generation, short-read sequencing technologies have enabled high-depth mtDNA analysis(3) and are currently considered the gold standard for heteroplasmy detection(7). However, these approaches rely on read lengths of ∼50-300bps, which can lead to uneven coverage, mapping biases, vulnerability towards NUMTs contamination, and limit the direct resolution of full-length mtDNA haplotypes. Additionally, longer read lengths can facilitate the assessment of structural variants, as reads can span larger genetic events.

Long-read sequencing technologies have the potential to overcome these limitations by enabling full-length mtDNA sequencing. It has been demonstrated that Oxford Nanopore long-read sequencing can accurately detect mtDNA heteroplasmy at variant levels >5% and requires higher sequencing depth (8, 9). PacBio HiFi sequencing provides highly accurate long reads, making it an interesting tool for detecting low-level variants. Still, systematic benchmarking of PacBio long-read sequencing for accurate mtDNA heteroplasmy detection remains limited, particularly at low variant levels (<5%). Additionally, the deep sequencing of full-length mtDNA molecules has often been compromised by the use of multiple overlapping PCR amplicons(5, 7, 8).

In this study, we explored the performance of PacBio HiFi sequencing for detecting mtDNA heteroplasmy using a single full-length long-range PCR approach. Utilizing predefined haplotype mixtures, we assessed detection accuracy across low variant levels. Finally, we applied this approach to investigate longitudinal heteroplasmic dynamics in fibroblast passages from LRRK2 p.Gly2019Ser variant carriers, affected by PD or unaffected.

## Methods

### Study design and sample overview

To evaluate the applicability of PacBio HiFi sequencing for detecting mtDNA heteroplasmy across the full mitochondrial genome, we used a two-part study design consisting of a mixture-based validation experiment and a biological application (**Figure 1**).

**Figure 1.**
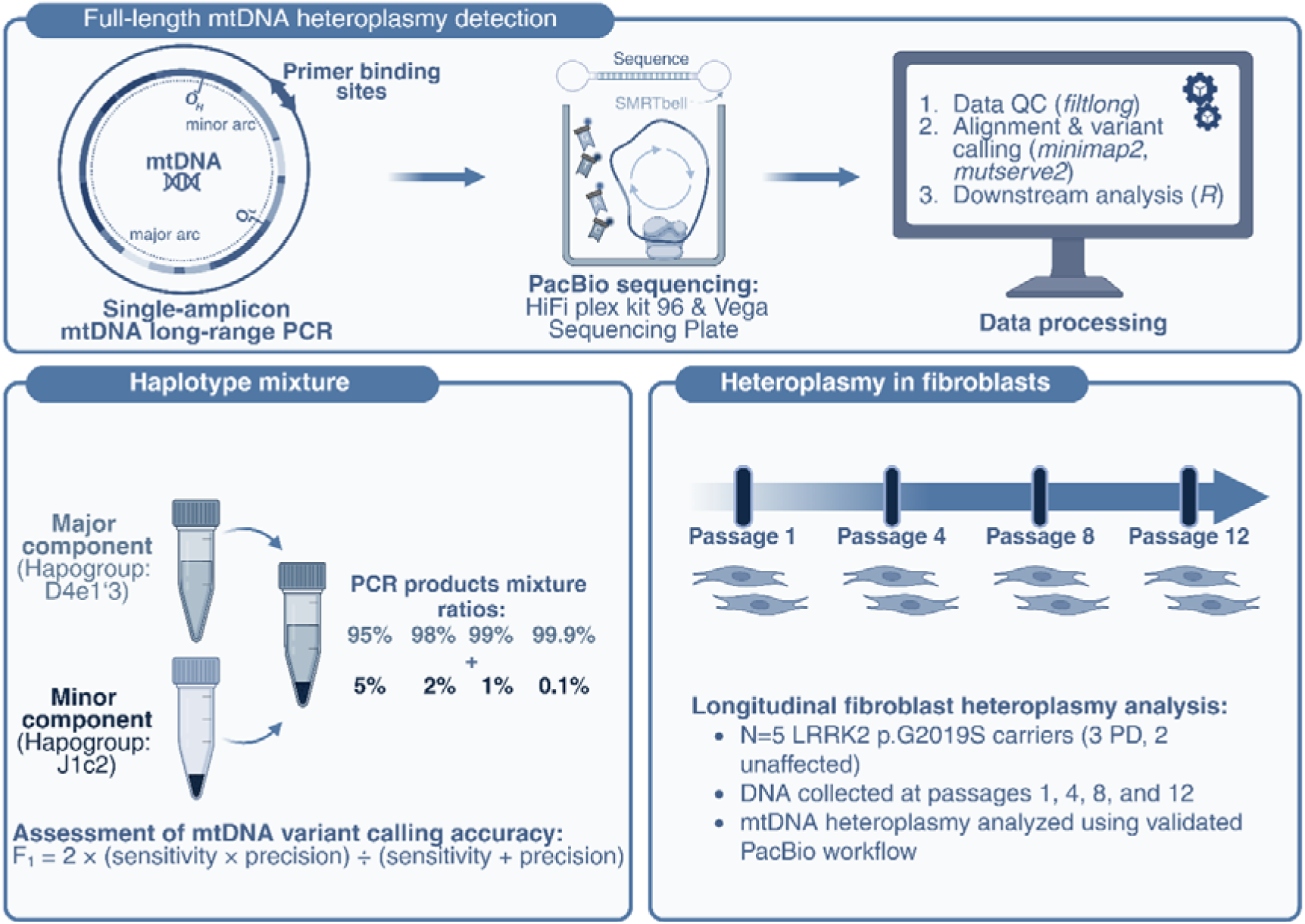
Study overview and workflow for full-length mtDNA heteroplasmy detection using PacBio HiFi sequencing. A single long-range PCR amplicon spanning the entire mitochondrial genome was generated and sequenced using the PacBio HiFi platform. Sequencing reads were subjected to quality control, alignment, and heteroplasmy detection using *minimap2* and *Mutserve2*, followed by downstream statistical analyses in R. To validate heteroplasmy detection accuracy, predefined mixtures of two mtDNA haplotypes were generated at ratios ranging from 5% to 0.1% and evaluated using the F1 score. The validated workflow was subsequently applied to investigate longitudinal mtDNA heteroplasmy dynamics across fibroblast passages (1, 4, 8, and 12) derived from five LRRK2 p.G2019S variant carriers.

First, predefined mixtures of two mtDNA haplotypes were generated to benchmark heteroplasmy detection accuracy across a range of low-frequency variant levels. As previously described, the DNA used in the mixture model was extracted from blood with the Blood and Cell Culture DNA Midi kit (Qiagen), and the selected samples were from participants of European ancestry(8) (**Supplementary Table 1**). These mixtures were prepared at defined proportions ranging from 95% (major component, haplogroup D4e1’3) plus 5% (minor component, haplogroup J1c2) to 99.9% (major component) plus 0.1% (minor component). The samples were selected based on their haplotypes, containing both shared variants and variants that differ between the two haplogroups. The samples had previously been characterized using Illumina NextSeq sequencing as a gold-standard method, allowing us to assess the accuracy of our workflow by comparing expected and observed variants, particularly those from the minor component that mimic mitochondrial heteroplasmy(8).

Second, the validated sequencing workflow was applied to investigate longitudinal heteroplasmic changes across time. We used fibroblast cell lines derived from five LRRK2 p.Gly2019Ser variant carriers (n=3 affected with PD and n=2 unaffected carriers, **Supplementary Table 1**). The rationale for selecting *LRRK2*-related cellular models is based on previous reports on mitochondrial mechanisms in LRRK2 penetrance(10–12). For this purpose, mtDNA was extracted from fibroblast cultures at passages 1, 4, 8, and 12 to assess changes in heteroplasmic variant load and heteroplasmy levels across cell passages longitudinally. In both experimental settings, deep mtDNA PacBio sequencing was performed using a single long-range PCR amplicon spanning the entire mitochondrial genome.

Written informed consent was obtained from all individuals, and the study was conducted according to the guidelines of the Declaration of Helsinki, and approved by the Ethics Committees of the University of Lübeck, Germany (protocol code 16-039, date of approval 27 September 2019).

### Long-Range PCR Amplification of mtDNA

One single amplicon spanning the complete mtDNA was prepared with long-range PCRs. The primer set binds within the minor arc of the mtDNA (chrM:2120F: 51-GGA CAC TAG GAA AAA ACC TTG TAG AGA GAG −31 and chrM:2119R: 51-AAA GAG CTG TTC CTC TTT GGA CTA ACA −31)(13). We used the high-fidelity Platinum SuperFi II DNA Polymerase, and the details for the PCR mix and conditions are listed in **Supplementary Table 2**.

### Preparation of haplotype mixture

The DNA concentration of the resulting PCR products was measured using a Qubit fluorometer and subsequently normalized. To generate the haplotype mixtures, PCR products of the major component (sample B-28, haplogroup D4e113) were combined with PCR products of the minor component (sample L-2804, haplogroup J1c2) at predefined proportions with minor mixture components of 5%, 2%, 1%, and 0.1% (**Supplementary Table 3**).

### LRRK2 variant carrier-derived fibroblast cultures

We included five fibroblast cultures from carriers of the LRRK2 p.Gly2019Ser variant, and all cultures were frozen at passage 3 prior to analysis. The cells stored in liquid nitrogen were thawed and cultured in T75 cell culture flasks using DMEM supplemented with 1% Penicillin-Streptomycin and 10% FBS. After reaching initial confluency, the cells were split into a new T75 cell culture flask every 7 days. At passages 1, 4, 8, and 12, half of the cells were collected in a 1.5ml Eppendorf Tube. The aliquot was centrifuged at 1000xg for 5 minutes at 4°C, and the supernatant was removed. The pellet was then washed once in DPBS, and after another centrifugation, the DPBS was removed, and the pellets were stored at −80°C until DNA extraction.

We extracted DNA longitudinally across passages. Specifically, we assessed mtDNA heteroplasmy using DNA extracted from passages 1 (day 0), 4 (day 28), 8 (day 56), and 12 (day 84). The DNA was extracted using the AutoGen XTRACT 16+ XK401 Kit for cultured cells, and the DNA quality was measured with the Qubit fluorometer and the Nanodrop. Subsequently, the mtDNA long-range PCR was performed for all fibroblast-derived DNA samples as described above.

### PacBio Library Preparation and Sequencing

All four mixture samples and all PCR products (DNA) derived from the fibroblasts were barcoded and sequenced together. We used 480ng of PCR-amplified DNA as the input for the library preparation, starting with the end repair, following the manufacturer’s instructions of the HiFi plex kit 96 (103-381-300) with the SMRTbell adapter index plate 96 A (102-009-200) and the Vega polymerase kit (103-426-500). The sequencing was performed on the PacBio Vega device with the Vega Sequencing Plate (103-274-300) and the Vega SMRT cell tray (103-406-700).

### Sequencing data processing

PacBio sequencing data generated on the Vega system were obtained as unmapped BAM files. Approximately 95% of the sequenced reads were classified as HiFi reads (base quality≥Q30). Reads were filtered using *Filtlong* (v0.2.1) (https://github.com/rrwick/Filtlong) to retain sequences with lengths between 15 kb and 18 kb. Filtered reads were aligned to the mitochondrial reference genome (revised Cambridge Reference Sequence, rCRS) using *Minimap2* (v2.22) (14). The resulting alignments were subsequently sorted and indexed using *Samtools* (v1.3.1) (15) and filtered for alignment lengths >1kb.

Our group previously benchmarked multiple alignment and variant-calling tools for assessing mtDNA heteroplasmy using Oxford Nanopore long-read sequencing, demonstrating that *Mutserve2* provided the highest variant-calling accuracy, while alignment tools had only a marginal impact on performance(8). Therefore, we used *Mutserve2* (v2.0.0-rc8)(16, 17), a tool specifically designed to detect heteroplasmic and homoplasmic mtDNA variants, for variant calling. Detected variants were annotated using the same tool, and potential sample contaminations was assessed using *Haplocheck* (v1.3.3) (18). To evaluate heteroplasmy detection accuracy across different sequencing depths in haplotype mixtures, we generated stepwise-downsampled subsets of the alignments using the *Samtools-s* option. In addition, we utilized *Himito*(19) a graph-based software tool for mtDNA analysis, to explore possible NUMTs contamination in the long-read data.

For the longitudinal fibroblast analysis, variant calling was performed using the complete sequencing dataset and the same data analysis workflow. Additionally, we used *Mitorsaw* (https://github.com/PacificBiosciences/mitorsaw) and *Sniffles2*(20) to call variants beyond SNVs in the fibroblasts. *Mitorsaw* is a variant caller specifically designed to call mtDNA variants from PacBio data, creating consensus sequences for each haplotype and performing an alignment to the looped version of the reference mitochondria, and we used the *--minimum-maf 0.01* parameter. *Sniffles2* is a variant caller for long-read sequencing data, and we used the *--mosaic* and *--mosaic-af-min 0.01* parameters.

### Statistical analysis

As our group previously described(8), to evaluate variant calling performance, detected variants were classified as true positives, false positives, or false negatives. True-positive variants were defined as those expected to be present in the predefined haplotype mixtures based on the previously generated Illumina NextSeq sequencing data used as the reference standard. False-positive variants were defined as variants detected by the PacBio workflow but discordant from the Illumina reference dataset, whereas false-negative variants were variants expected in the mixtures but not detected.

Based on these classifications, the F_1_ score was calculated as a combined measure of precision and sensitivity (F_1_ score = 2 × (precision × sensitivity) ÷ (precision + sensitivity)). For the calculation of the F_1_ score and the determination of false-positive calls, minimum heteroplasmy thresholds were applied to the *Mutserve2* output according to the expected mixture proportions: ≥0.025 for the 5% mixture, ≥0.010 for the 2% mixture, ≥0.005 for the 1% mixture, and ≥0.0005 for the 0.1% mixture. In addition, the number of false-positive variants was calculated to assess further variant calling performance. For the mixtures, we can differentiate between three different types of variants: 1) major variants (only present in the major component and expected at 95%-99.9%), 2) minor variants (only present in the minor component and expect at 5%-0.1%), and 3) common variants (present in both samples and expect at 100% at all mixture rations).

Lastly, changes in mtDNA variant load across fibroblast passages were assessed using a linear mixed-effects model. Passage number, age at extraction, and PD status were included as fixed effects, while the fibroblast cell line was modeled as a random effect. The analyses were performed in *R* (v.4.3.1) (21) with the *lme4* (v.1.1-34) (22) and *lmerTest* (v.3.1-3) (23) packages.

## Results

First, we evaluated the sequencing performance across all multiplexed samples sequenced for this study. PacBio HiFi sequencing generated high-quality full-length mtDNA reads across all haplotype mixtures and fibroblast samples. More than 95% of all sequenced DNA reads achieved a basecalling quality Phred score of at least 30, corresponding to an accuracy of 99.9%. Filtering for q-score, read and alignment length, ∼21% of the sequenced reads were discarded mostly due to the read length (**Supplementary Figure 1**). After applying the described filtering steps, the majority of the reads showed a read length of ∼16.5kb, representing to the full-length mtDNA amplicon (**Supplementary Table 4, Supplementary Figure 1**).

### High sequencing depth was obtained across all 24 samples, with an average coverage of 83,180X

(range: 64,035X-107,279X) after filtering for read and alignment lengths. Coverage was highly uniform across all positions of the mitochondrial genome, indicating even amplification and sequencing of the full-length amplicon (**Supplementary Figure 2A**, exemplary sample). We randomly subsampled 0.5% of the sequenced reads and observed similarly even coverage across the entire mitochondrial genome at an approximate sequencing depth of 320X (**Supplementary Figure 2B**).

### PacBio workflow is highly accurate for mtDNA variant detection in mixture models

To evaluate the accuracy of mtDNA heteroplasmy detection using the PacBio workflow, we analyzed predefined mixtures of two mtDNA haplotypes over a range of minor-haplotype fractions. Variant calling was performed using *Mutserve2* and compared with the expected variant profiles derived from the Illumina NextSeq reference data, as a gold-standard method. Across mixture levels from 5% to 1%, heteroplasmic variants were detected with perfect concordance between expected and observed variants, resulting in an F_1_ score of 1.0 (**Table 1**). Thus, we also detected no additional variants discordant with the Illumina reference in the 5%, 2%, or 1% mixtures.

**Table 1.**
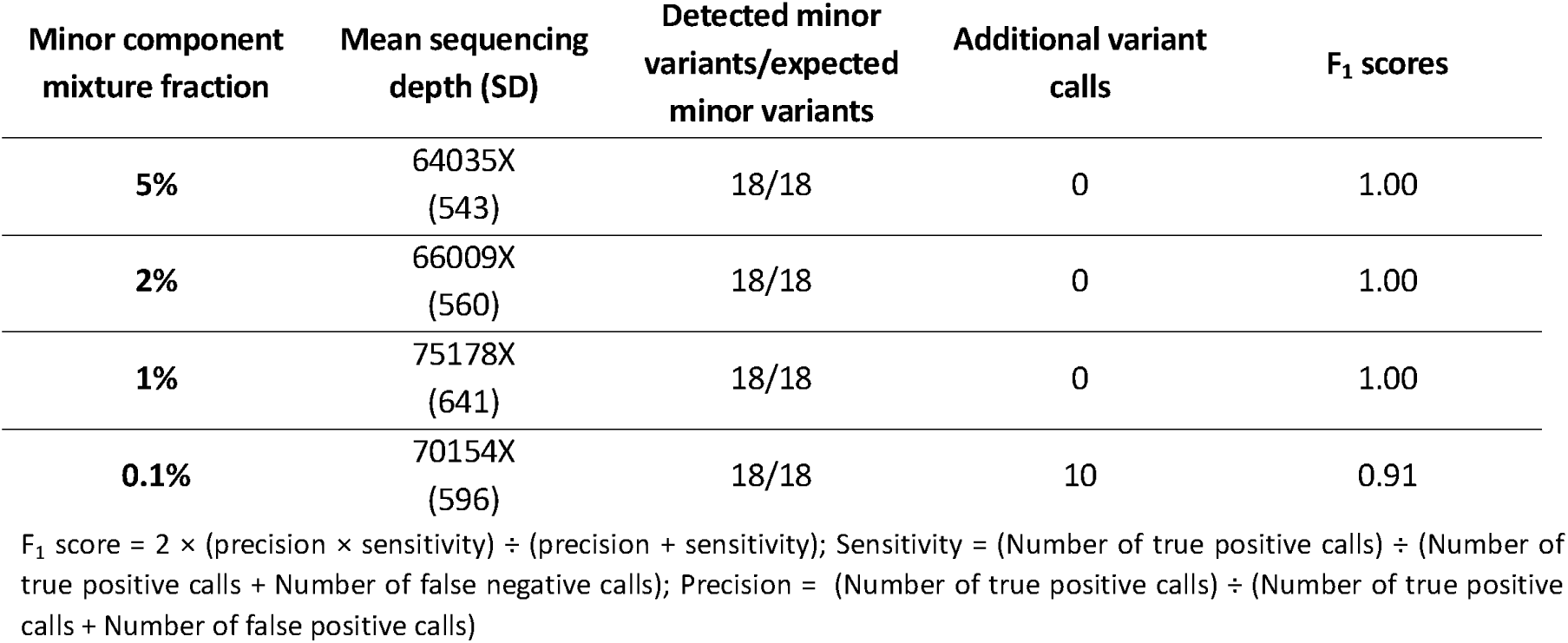
Summary of the coverage and accuracy measures detected from the mixture models.

To assess the detection limits of the workflow, we additionally included a mixture containing 0.1% of the minor component. Sensitivity remained high, with all minor variants detected at the 0.1% heteroplasmy level; however, 10 additional variants were observed, resulting in an F_1_ score of 0.91. Importantly, the detected variant levels of minor variants across all mixtures closely matched the expected levels based on the predefined mixture ratios, with minimal deviation between individual variants (**Figure 2A**).

**Figure 2.**
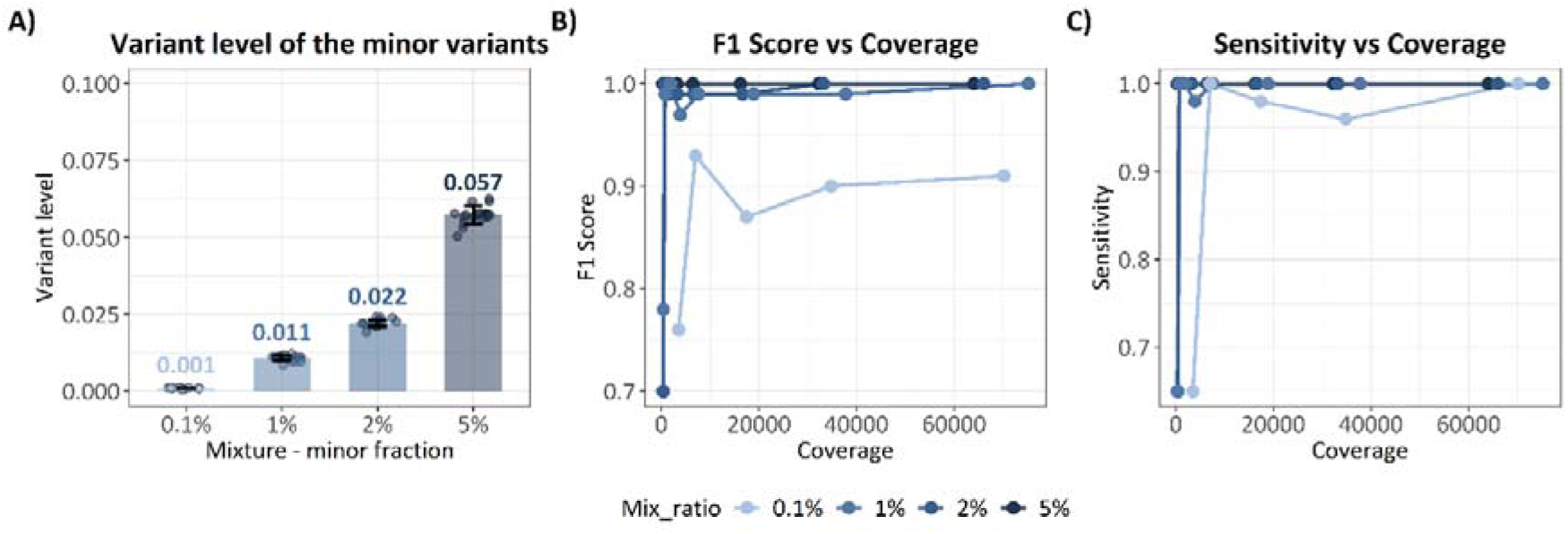
Validation of mtDNA heteroplasmy detection accuracy using predefined haplotype mixtures. **(A)** Detected heteroplasmy levels of the minor component across mixture ratios. Bars indicate mean values and standard deviation, and points represent individual measurements. **(B)** F_1_ score across sequencing coverage levels for mixtures containing minor component fractions of 0.1%, 1%, 2%, and 5%. **(C)** Sensitivity of variant detection across coverage levels for the same mixture ratios.

Given the high sequencing depth obtained across all multiplexed samples, we next evaluated variant calling accuracy and sensitivity at different sequencing depths by stepwise subsampling the data (**Figure 2B-C**). Accuracy and sensitivity remained very high at sequencing depths >1000X (F_1_ score range: 0.97-1.00, sensitivity range: 0.98-1.00, **Supplementary Table 5**) for mixture ratios ≥1%. The F_1_ score and sensitivity remained 1.00 across all tested sequencing depths for the 5% mixture, even at the lowest evaluated coverage of approximately 300X.

Lastly, we utilized *Himito* to explore the indication of NUMTs contamination in all four mixtures. Using default parameters of the toolkit, there was no evidence for any NUMTs contamination in the data set.

### No detectable systematic sequencing errors and minimal sequencing noise

Type I errors (i.e., variants discordant with the Illumina reference) were investigated following the removal of all expected variants and were observed exclusively in the 0.1% haplotype mixture. Further characterization using the reference genome GRCh38 showed that these variants were not due to systematic errors on specific sites and were partially detected in low-complexity regions, such as short homopolymers (**Supplementary Table 6**). Additionally, we used *Mitorsaw*, a haplotype-aware variant caller, to assess the context of these additional variant calls in the 0.1% mixture. Interestingly, only two out of the ten additional variants discordant from the Illumina reference were also detected with *Mitorsaw* at a similar variant level as *Mutserve2*, and they did not cluster on the same haplotype (i.e., chrM:299A and chrM:1419A).

Common variants shared between major and minor components are expected to be consistently detected at a homoplasmic level of 1. As such, they serve as internal controls for assessing technical noise within the dataset, where any deviation from 1 would reflect technical or stochastic variation. These variants were further evaluated within the 5% haplotype mixture to validate the accuracy of the sequencing workflow (**Supplementary Table 7**). Across the full data set (∼64,000 sequencing depth), variant levels remained at 1, and a high level of concordance was maintained after subsampling to approximately 300X depth, with no meaningful deviation observed (variant level ≥0.997).

### Mitochondrial heteroplasmic variant load is associated with PD status in LRRK2 variant carriers

To investigate longitudinal mtDNA heteroplasmy dynamics, we applied the validated PacBio workflow to fibroblast cell lines derived from five LRRK2 p.Gly2019Ser variant carriers, three PD patients, and two unaffected carriers. Heteroplasmic variants were assessed across fibroblast passages 1 (day 0), 4 (day 28), 8 (day 56), and 12 (day 84); all cell cultures were frozen at passage 3 prior to our analysis. Based on our validation using the mixture model, we restricted the longitudinal analysis to heteroplasmic variants with heteroplasmy levels >1%. Across the 12 passages, we observed a decrease in the number of detected heteroplasmic mtDNA variants (**Figure 3**). To quantify this trend, we applied a linear mixed-effects model including passage number as the time variable, affection status, age as fixed effects, and the five individual cell lines as a random effect (**Table 2**). The model confirmed that over time and across passages, there were fewer heteroplasmic variants (passage 12 vs passage 1: β=-3.2, SE=1.3, p=0.026). Notably, mtDNA heteroplasmic variant load was higher in fibroblast lines derived from affected LRRK2 variant carriers compared to unaffected carriers (β=5.0, SE=1.0, p=1.0×10^-4^). In addition, mtDNA heteroplasmy was associated with older age at sample collection (β=1.5, SE=0.3, p=0.001).

**Figure 3.**
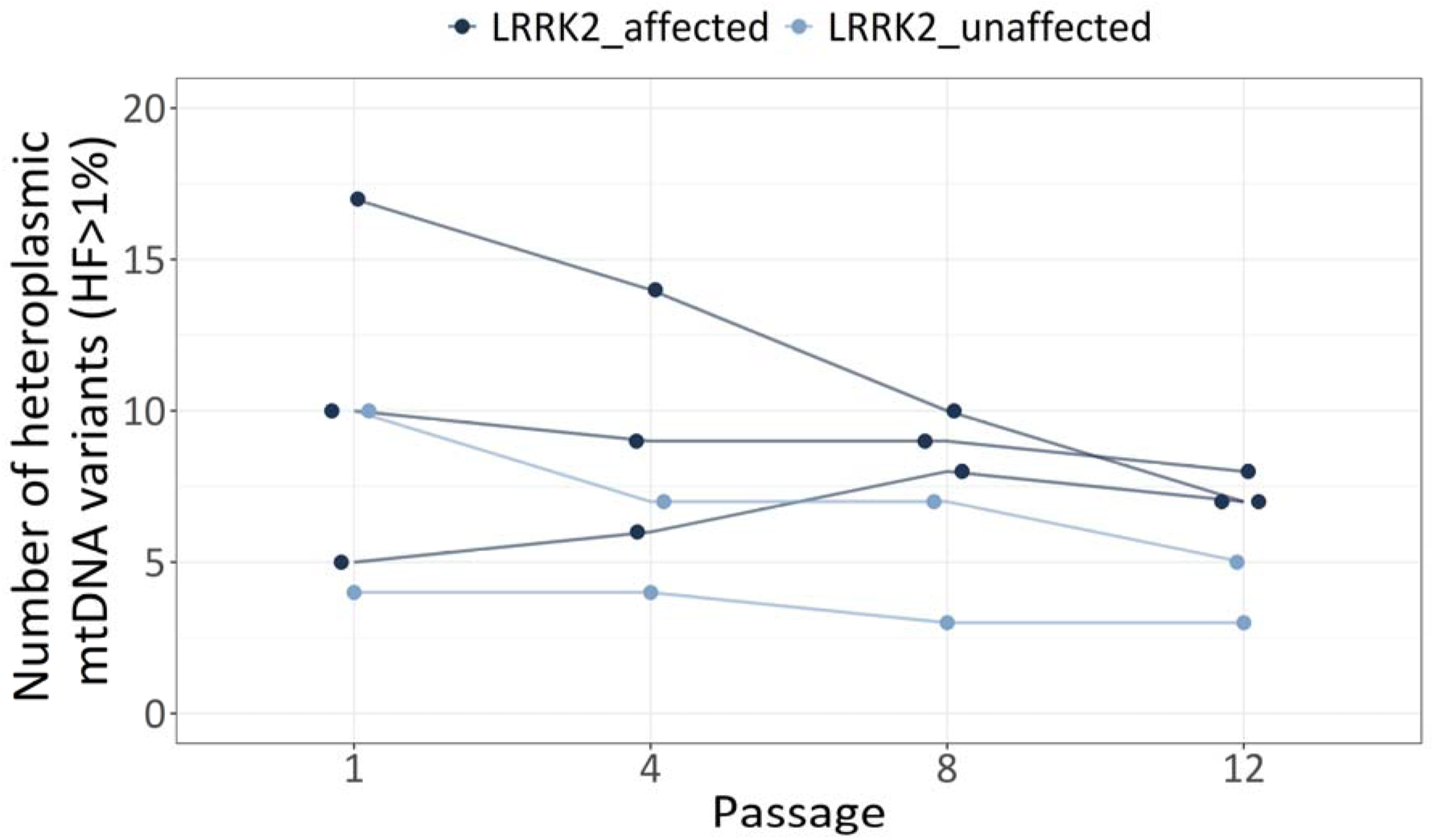
Longitudinal changes in heteroplasmic variant load across fibroblast passages in LRRK2 variant carriers. The number of mtDNA variants with heteroplasmy levels >1% was quantified across fibroblast passages (1 (day 0), 4 (day 28), 8 (day 56), and 12 (day 84)) derived from five LRRK2 p.G2019S variant carriers, including three individuals affected with Parkinson’s disease (dark blue) and two unaffected carriers (light blue). Each line represents an individual cell line.

**Table 2.** Linear mixed-effects model assessing factors associated with the number of mtDNA heteroplasmic variants.

|  | Estimate | Standard error | p-value |
| --- | --- | --- | --- |
| **Passage 4** | -1.2 | 1.3 | 0.367 |
| **Passage 8** | -1.8 | 1.3 | 0.184 |
| **Passage 12** | -3.2 | 1.3 | 0.026 |
| **Affection status: *LRRK2*-PD** | 5.0 | 1.0 | $1.0 \times 10^{-4}$ |
| **Age at sample collection** | 1.5 | 0.3 | 0.001 |

PD=Parkinson's disease, Baseline category: Passage=1, Affection status=*LRRK2* unaffected
R-formula: lmer (Overall.Heteroplasmies ~ Passage + Affection status + age + (1 | Sample ID)); Models include fibroblast cell line as a random effect.

As dynamic changes in mtDNA heteroplasmy were observed across fibroblast passages, variant-specific profiles were evaluated by examining mitochondrial genomic positions and heteroplasmy levels at each passage. This analysis revealed heterogeneous dynamics of specific mtDNA variants over time. While some variants gradually decreased in heteroplasmy levels across passages, others showed increasing levels, indicating variant-specific trajectories during fibroblast culture. For example, in the fibroblast line L-15695 derived from an affected *LRRK2* carrier, we observed that the heteroplasmic variant levels of the variants at chrM:7642A (MT-CO2, synonymous) and chrM:14118G (MT-ND5, synonymous) decreased over time. In contrast, the variant at chrM:4818A (MT-ND2, missense p.E117K) showed a gradual increase in heteroplasmy level across passages (**Figure 4**). Overall, heteroplasmic variant profiles were highly heterogeneous across samples and passages, with no consistent pattern observed between affected and unaffected LRRK2 p.Gly2019Ser carriers (**Supplementary Figure 3**).

**Figure 4.**
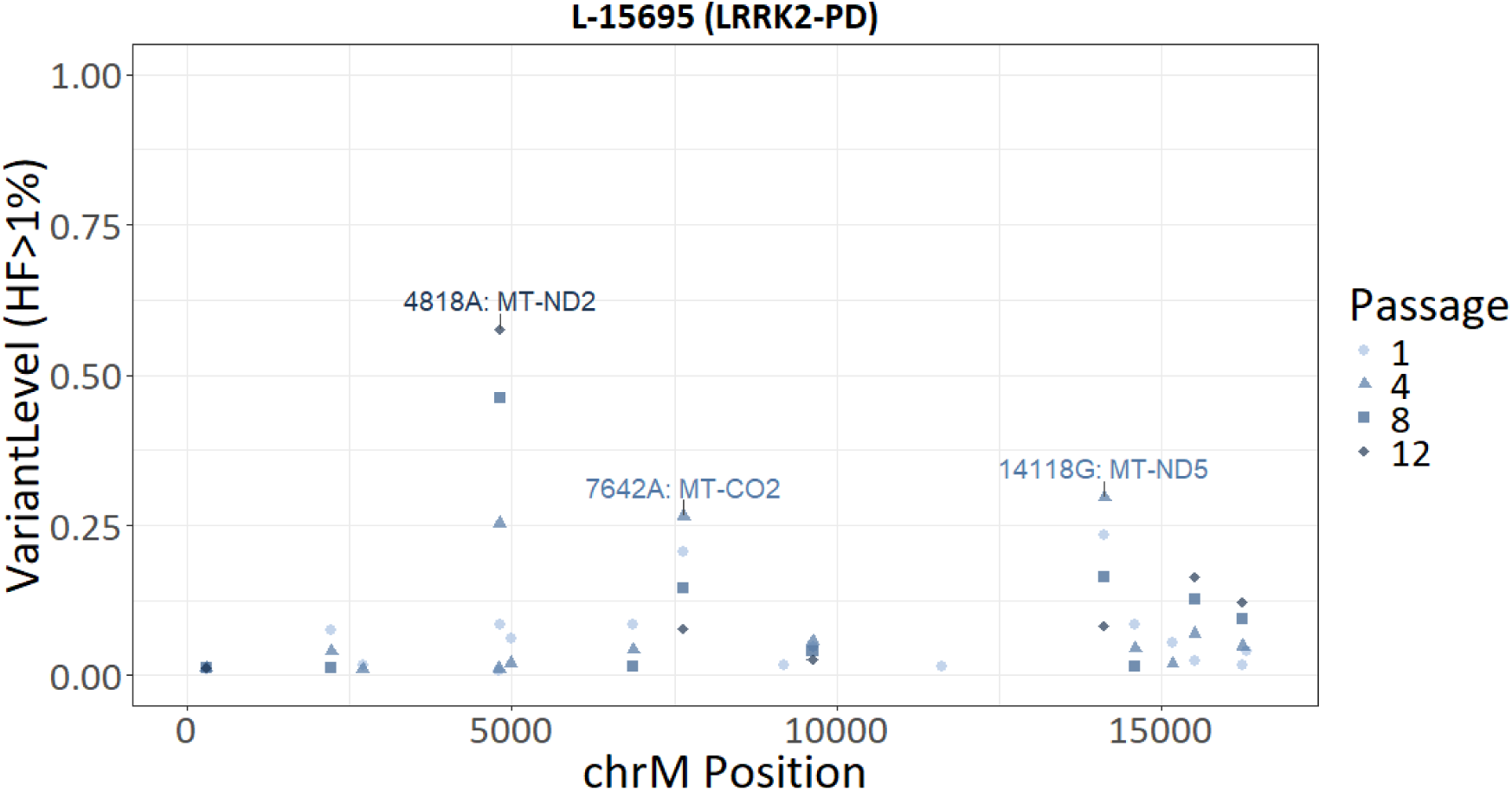
Variant-specific heteroplasmy dynamics across fibroblast passages in an LRRK2 p.Gly2019Ser carrier with Parkinson’s disease (PD). Heteroplasmy levels of mtDNA variants (>1%) plotted across mitochondrial genome positions (chrM) for fibroblast cell line L-15695 derived from an affected LRRK2 variant carrier. The different symbols and colors represent heteroplasmy levels detected at fibroblast passages 1, 4, 8, and 12. Several variants exhibit distinct variant level changes across passages, including variants located in the genes MT-ND2 *(chrM:4818A),* MT-CO2 *(chrM:7642A), and MT-ND5 (chrM:14118G)*.

Utilizing *Mitorsaw* and *Sniffles2* to explore structural variants like deletions, we did not detect any at an allele frequency >1%.

## Discussion

In this study, we developed and validated a PacBio HiFi-based workflow for accurate detection of mitochondrial DNA heteroplasmy using a single full-length mtDNA amplicon. Utilizing a mitochondrial haplotype mixture model, we demonstrate very high accuracy, with 100% concordance between expected and detected variants down to the 1% variant level. Across all multiplexed samples, PacBio HiFi sequencing generated high-quality full-length reads with consistent read lengths and uniformly high coverage across the mitochondrial genome.

Sensitivity and accuracy remained high even at reduced, subsampled sequencing depth, suggesting that large numbers of samples can be multiplexed without losing heteroplasmy detection accuracy. In detail, given the sequencing depth obtained in this study, multiplexing all 230 available barcodes would still yield approximately 13,000X mtDNA coverage per sample, enabling accurate detection of mitochondrial heteroplasmy levels at 1%. We also tested the detection limits of the targeted PacBio mtDNA workflow by including a mixture containing 0.1% of the minor component. Although sensitivity remained high and all minor variants were detected at the expected variant-level frequency of 0.001, the assessment of such low-level variants was affected by variants discordant with the Illumina reference, reducing overall calculated accuracy. In line with previous findings^8^, these discordant variants may reflect residual sequencing and alignment-related noise, particularly at very low variant levels (0.1%). Assessment of common variants further confirmed stable detection at homoplasmic levels (variant level ∼1.00), indicating low technical noise. It is essential to note that the use of the Illumina dataset as a gold-standard reference may overestimate the number of discordant variants at very low variant levels, as orthogonal validation (e.g., sequencing of unmixed samples) was not performed in this study.

To our knowledge, we are the first to demonstrate the applicability of PacBio HiFi sequencing for highly accurate detection of mitochondrial heteroplasmy by integrating real sequencing data with a haplotype mixture model. Previous studies have explored the accuracy of mtDNA heteroplasmic variant calling using long-read sequencing technologies, primarily ONT. Reliable detection of relatively high heteroplasmy levels >12%(24) or >10%(25) were reported, but partially with lower sequencing depth(24).

In our previous work, we evaluated the accuracy of mtDNA heteroplasmy variant calling using deep, targeted mtDNA ONT sequencing across two overlapping PCR amplicons. In contrast, we demonstrated reliable mtDNA heteroplasmy variant calling >5% variant levels(8). However, the overall accuracy for very low-frequency variants below 5% remained limited with ONT sequencing. Herein, we demonstrated that our established PacBio workflow has the potential to close this gap and detect mitochondrial heteroplasmy down to 1% or even further.

The precise assessment of low-frequency variants is important for various research fields, including cancer research and the investigation of mosaic somatic variants and of course mitochondrial heteroplasmy(2, 5, 26, 27). More broadly, genome-wide benchmarking studies of somatic variant detection have reported lower F_1_ scores and recall-rates/sensitivity with lower whole-genome sequencing depths (∼40X-60X)(28–30). This highlights the advantage of the targeted high-depth sequencing approaches for detecting rare variants in the mtDNA.

Some limitations of the presented workflow are important to note. The use of long-range PCR introduces the risk of primer dropout if polymorphisms occur within the primer binding sites, which may lead to complete amplification failure. Most prominently Haplogroup: D1, H1n, M24b, U3, U7, and T2a, which all have known polymorphisms at chrM:2090-2150. PCR amplification can also introduce low variant-level polymerase errors that might have been missed in the present benchmarking. In addition, PCR-based enrichment removes native DNA methylation, and thus, analysis of mitochondrial epigenetic architecture is not possible with our workflow. Furthermore, our validation focused on single-nucleotide variant detection. The performance of this workflow for structural variant detection was not systematically evaluated. However, the use of a single full-length mtDNA amplicon may, in principle, facilitate structural variant detection compared to multi-amplicon approaches, provided that variants do not disrupt the primer-binding sites. While we explored the presence of structural variants, such as deletions, none were detected in the analyzed samples. This likely reflects the absence of structural variants above the applied detection threshold (>1%) rather than a limitation of the approach. Consistent with this, the read length distribution of the unfiltered sequencing data did not show an enrichment of shorter reads, as would be expected in the presence of deletion-containing molecules.

Finally, we used the newly established and validated PacBio HiFi workflow to assess mtDNA heteroplasmy over time in fibroblasts derived from carriers of the LRRK2 p.Gly2019Ser variant. Based on the results from the mixture validation, we focused on heteroplasmic variants with variant levels >1%.

Overall, we observed a decrease in the mitochondrial heteroplasmic variant load across all cell lines over time. Indeed, evaluating the individual positions and levels of detected heteroplasmic variants showed that certain variants exhibit a general decrease in their levels over time. There were even variants that were detected in passages 1 to 8 but no longer in passage 12 (**Figure 4**). However, other variants exhibit a gradual increase in variant levels across passages (e.g. chrM:4818A in *MT-ND2*, **Figure 4**). Taken together, this may suggest that selective pressures or clonal expansion processes are shaping mitochondrial heteroplasmy during fibroblast passaging. A recent study investigated mtDNA heteroplasmy dynamics with a single-cell approach at different cell culture conditions. It was demonstrated that heteroplasmy levels in dividing cells are primarily shaped by selection rather than random drift(31). Furthermore, deleterious mtDNA variants can be selectively removed or not, depending on improvements in cell fitness under different cell culture conditions(31). Similarly, a study investigating mtDNA heteroplasmy during iPSC reprogramming and passaging reported no evidence for the emergence of novel heteroplasmic variants but observed that selection during passaging contributes to shaping heteroplasmy(32). It is important to note that we did not detect a shared pattern of dynamically changing variants across all analyzed samples, indicating substantial inter-individual variability in heteroplasmy trajectories.

We investigated heteroplasmy over time in fibroblasts derived from three *LRRK2* variant carriers affected with PD and two unaffected carriers, and the heteroplasmic variant load was higher in fibroblasts from affected than from unaffected carriers. Mitochondrial dysfunction is a key player in PD pathogenesis, including impairment of mtDNA integrity(33). Thus, our findings add to the growing body of evidence linking increased mtDNA heteroplasmy or damage to LRRK2-related PD, as highlighted by previous studies(10–12). Notably, somatic major arc deletions have been reported to be increased in fibroblast cultures derived from LRRK2-PD patients and to discriminate affected from unaffected variant carriers(12). To our knowledge, the work presented here is one of the first studies to longitudinally assess mtDNA heteroplasmy dynamics across fibroblast passages using high-accuracy long-read sequencing in the context of *LRRK2-*related PD, providing new insights into variant-specific trajectories and the role of selection in shaping mitochondrial variation over time.

Future studies should aim to investigate the landscape of mtDNA heteroplasmy in larger cohorts to thoroughly explore mitochondrial heteroplasmy in *LRRK2*-related PD or other PD subtypes, implementing long-read sequencing approaches.

### Conclusion

PacBio HiFi sequencing combined with a single-amplicon strategy facilitates accurate full-length mtDNA heteroplasmy detection and longitudinal analysis. Our benchmarking demonstrates highly accurate detection of heteroplasmic variants down to 1%, while detection at 0.1% retained high sensitivity but was affected by variants discordant with the reference set. Application of this workflow to *LRRK2* p.Gly2019Ser variant carrier fibroblast models revealed dynamic, variant-specific changes in heteroplasmy over time, and an association between higher heteroplasmic variant load and *LRRK2*-PD. Together, this approach provides a powerful tool to study mitochondrial heteroplasmy and its role in aging and disease.

## Declarations

### Ethics approval and consent to participate

Written informed consent was obtained from all individuals, and the study was conducted according to the guidelines of the Declaration of Helsinki, and approved by the Ethics Committees of the University of Lübeck, Germany (protocol code 16-039, date of approval 27 September 2019).

### Consent for publication

Not applicable

### Availability of data and materials

The generated data for this study will be made available for qualified researchers upon reasonable request.

### Authors’ contributions

TL, HW, and JT conceptualized the study. TL, MMB, and JT wrote the original draft of the manuscript. SS, CM, and PS provided the fibroblast cell lines, performed cell culture experiments, and generated the sequencing data. TL performed the visualization and generated the figures. TL, MMB, and HW performed the data analysis. HW and PM analyzed the Illumina reference sequencing data. JT, AG and CK provided resources and scientific supervision. All authors reviewed, edited, and approved the final manuscript.

## Acknowledgment

We gratefully acknowledge Dr. Tatjana Usnich and Dr. Vera Tadic for their contribution to the collection and provision of fibroblast samples used in this work.

## List of Abbreviations

AF: Allele Frequency
BAM: Binary Alignment Map
bp: Base Pairs
chrM: Mitochondrial Chromosome
DNA: Deoxyribonucleic Acid
F1: score Harmonic Mean of Precision and Sensitivity
HiFi: High-Fidelity
iPSC: Induced Pluripotent Stem Cell
kb: Kilobase
LRRK2: Leucine-Rich Repeat Kinase 2
mtDNA: Mitochondrial DNA
ONT: Oxford Nanopore Technology
PCR: Polymerase Chain Reaction
PD: Parkinson’s Disease
Q-score: Phred Quality Score
rCRS: Revised Cambridge Reference Sequence
SE: Standard Error
SNV: Single Nucleotide Variant
SV: Structural Variant
VAF: Variant Allele Frequency
X: Sequencing Coverage Depth

## References

1. Pinto M, Moraes CT. Mechanisms linking mtDNA damage and aging. Free Radic Biol Med. 2015;85:250–8.

2. Stewart JB, Chinnery PF. The dynamics of mitochondrial DNA heteroplasmy: implications for human health and disease. Nat Rev Genet. 2015;16(9):530–42.

3. Gupta R, Kanai M, Durham TJ, Tsuo K, McCoy JG, Kotrys AV, et al. Nuclear genetic control of mtDNA copy number and heteroplasmy in humans. Nature. 2023;620(7975):839–48.

4. Dulovic-Mahlow M, Konig IR, Trinh J, Diaw SH, Urban PP, Knappe E, et al. Discordant Monozygotic Parkinson Disease Twins: Role of Mitochondrial Integrity. Ann Neurol. 2021;89(1):158–64.

5. Trinh J, Hicks AA, Konig IR, Delcambre S, Luth T, Schaake S, et al. Mitochondrial DNA heteroplasmy distinguishes disease manifestation in PINK1/PRKN-linked Parkinson’s disease. Brain. 2023;146(7):2753–65.

6. Fang Z, Barbosa C, Marinho D, Peixoto R, Ferreira IL, Laranjinha J, et al. Linking mitochondrial DNA release to neurodegeneration and cognitive decline. Ageing Res Rev. 2026;117:103062.

7. Fazzini F, Fendt L, Schonherr S, Forer L, Schopf B, Streiter G, et al. Analyzing Low-Level mtDNA Heteroplasmy-Pitfalls and Challenges from Bench to Benchmarking. Int J Mol Sci. 2021;22(2).

8. Luth T, Schaake S, Grunewald A, May P, Trinh J, Weissensteiner H. Benchmarking Low-Frequency Variant Calling With Long-Read Data on Mitochondrial DNA. Front Genet. 2022;13:887644.

9. Dobner J, Nguyen T, Pavez-Giani MG, Cyganek L, Distelmaier F, Krutmann J, et al. mtDNA analysis using Mitopore. Mol Ther Methods Clin Dev. 2024;32(2):101231.

10. Howlett EH, Jensen N, Belmonte F, Zafar F, Hu X, Kluss J, et al. LRRK2 G2019S-induced mitochondrial DNA damage is LRRK2 kinase dependent and inhibition restores mtDNA integrity in Parkinson’s disease. Hum Mol Genet. 2017;26(22):4340–51.

11. Qi R, Sammler E, Gonzalez-Hunt CP, Barraza I, Pena N, Rouanet JP, et al. A blood-based marker of mitochondrial DNA damage in Parkinson’s disease. Sci Transl Med. 2023;15(711):eabo1557.

12. Ouzren N, Delcambre S, Ghelfi J, Seibler P, Farrer MJ, Konig IR, et al. Mitochondrial DNA Deletions Discriminate Affected from Unaffected LRRK2 Mutation Carriers. Ann Neurol. 2019;86(2):324–6.

13. Keraite I, Becker P, Canevazzi D, Frias-Lopez C, Dabad M, Tonda-Hernandez R, et al. A method for multiplexed full-length single-molecule sequencing of the human mitochondrial genome. Nat Commun. 2022;13(1):5902.

14. Li H. New strategies to improve minimap2 alignment accuracy. Bioinformatics. 2021;37(23):4572–4.

15. Danecek P, Bonfield JK, Liddle J, Marshall J, Ohan V, Pollard MO, et al. Twelve years of SAMtools and BCFtools. Gigascience. 2021;10(2).

16. Weissensteiner H, Forer L, Fuchsberger C, Schopf B, Kloss-Brandstatter A, Specht G, et al. mtDNA-Server: next-generation sequencing data analysis of human mitochondrial DNA in the cloud. Nucleic Acids Res. 2016;44(W1):W64–9.

17. Weissensteiner H, Forer L, Kronenberg F, Schonherr S. mtDNA-Server 2: advancing mitochondrial DNA analysis through highly parallelized data processing and interactive analytics. Nucleic Acids Res. 2024;52(W1):W102–W7.

18. Weissensteiner H, Forer L, Fendt L, Kheirkhah A, Salas A, Kronenberg F, et al. Contamination detection in sequencing studies using the mitochondrial phylogeny. Genome Res. 2021;31(2):309–16.

19. Su H, Huang Y, Durham T, Kong N, Casey E, Benjamin D, et al. Himito: a Graph-based Toolkit for Mitochondrial Genome Analysis using Long Reads. bioRxiv. 2025:2025.11.03.686348.

20. Smolka M, Paulin LF, Grochowski CM, Horner DW, Mahmoud M, Behera S, et al. Detection of mosaic and population-level structural variants with Sniffles2. Nat Biotechnol. 2024;42(10):1571–80.

21. R Core Team. R: A Language and Environment for Statistical Computing. R version 4.3.1 (2023-06-16 ucrt) ed2023.

22. Bates D, Mächler M, Bolker B, Walker S. Fitting Linear Mixed-Effects Models Using lme4. Journal of Statistical Software. 2015;67(1):1–48.

23. Kuznetsova A, Brockhoff PB, Christensen RHB. lmerTest Package: Tests in Linear Mixed Effects Models. Journal of Statistical Software. 2017;82(13):1–26.

24. Slapnik B, Sket R, Crepinsek K, Tesovnik T, Bizjan BJ, Kovac J. The quality and detection limits of mitochondrial heteroplasmy by long read nanopore sequencing. Sci Rep. 2024;14(1):26778.

25. Song H, Liu J, Yang F, Du W, Qin L, Su Y, et al. A long-amplicon nanopore sequencing and analysis method for human whole mitochondrial genome. Forensic Sci Int. 2025;377:112668.

26. Watson IR, Takahashi K, Futreal PA, Chin L. Emerging patterns of somatic mutations in cancer. Nat Rev Genet. 2013;14(10):703–18.

27. Trinh J, Luth T, Schaake S, Laabs BH, Schluter K, Labeta J, et al. Mosaic divergent repeat interruptions in XDP influence repeat stability and disease onset. Brain. 2023;146(3):1075–82.

28. Zhang Y, English AC, Paulin LF, Grochowski CM, Maheshwari S, Mack T, et al. Comprehensive benchmarking of somatic structural variant detection at ultra-low allele fractions. bioRxiv. 2025.

29. Park J, Cook DE, Chang PC, Kolesnikov A, Brambrink L, Mier JC, et al. Accurate somatic small variant discovery for multiple sequencing technologies with DeepSomatic. Nat Biotechnol. 2025.

30. Somatic Mosaicism across Human Tissues N, Abyzov A. Comprehensive benchmarking of somatic mutation detection by the SMaHT Network. bioRxiv. 2025.

31. Kotrys AV, Durham TJ, Guo XA, Vantaku VR, Parangi S, Mootha VK. Single-cell analysis reveals context-dependent, cell-level selection of mtDNA. Nature. 2024;629(8011):458–66.

32. Kosanke M, Davenport C, Szepes M, Wiehlmann L, Kohrn T, Dorda M, et al. iPSC culture expansion selects against putatively actionable mutations in the mitochondrial genome. Stem Cell Reports. 2021;16(10):2488–502.

33. Lang M, Grunewald A, Pramstaller PP, Hicks AA, Pichler I. A genome on shaky ground: exploring the impact of mitochondrial DNA integrity on Parkinson’s disease by highlighting the use of cybrid models. Cell Mol Life Sci. 2022;79(5):283.

