## Supplementary for "Long-read sequencing enables high-accuracy mitochondrial heteroplasmy detection in Parkinson’s disease"

**Supplementary material:**

**Supplementary Table 1. Overview of samples used for the mixture model and in the fibroblast assessment.**

| **Mixture model** | | | | |
| --- | --- | --- | --- | --- |
| **ID** | **Age at sample collection range**  **(years)** | **Ancestry & Country** | **Mitochondrial haplogroup** | |
| **B-28** | 30-40 | European / Italy | D4e1’3 | |
| **L-2804** | 70-80 | European / Germany | J1c2 | |
| **Fibroblast cell lines derived from LRRK2 p.Gly2019Ser variant carriers** | | | | |
| **ID** | **Age at sample collection range**  **(years)** | **Ancestry & Country** | **Affection status** | **Mitochondrial haplogroup** |
| **L-15695** | 50-60 | European / Germany | Parkinson’s disease | H5 |
| **L-20393** | 50-60 | European / Germany | Parkinson’s disease | H5a1b |
| **L-15935** | 50-60 | European / Germany | Parkinson’s disease | T1a1k |
| **L-3086** | 50-60 | European / Germany | Unaffected | J1c2p |
| **L-3085** | 50-60 | European / Germany | Unaffected | J1c2p |

**Supplementary Table 2. Mitochondrial DNA Long-range PCR mix for one sample and cycling conditions for amplifying the single mitochondrial amplicon.**

| **PCR master-mix** | |
| --- | --- |
| **Reagent** | **1x [µl]** |
| SuperFi II OCR MM | 12.5 |
| 10µM Primer F | 1.25 |
| 10µM Primer R | 1.25 |
| DNA (50ng) | X |
| Nuclease-free water | 10.0-X |
| ∑ | 25.0 |
| **Cycler conditions** | |
| **Temperature and duration** | **Process and cycles** |
| 94°C for 1:00 | Initial denaturation |
| 98°C for 0:10 | 30 cycles |
| 68°C for 16:00 |  |
| 72°C for 10:00 | Final extension |
| 4°C forever | ---- |

**Supplementary Table 3. Preparation of the mixtures of the PCR products at predefined 1%, 2%, 1% and 0.1% ratio.**

|  | | **Major component: D4e1’3** | **Minor component: J1c2** |
| --- | --- | --- | --- |
| **Volume of PCR product [µl]** | **0.1%** | 49.95 | 0.05 |
|  | **1%** | 49.5 | 0.5 |
|  | **2%** | 49.0 | 1.0 |
|  | **5%** | 47.5 | 2.5 |

**Supplementary Table 4. Overview of sequencing quality parameters across all multiplexed samples.**

| **Sample** | **Median Q score** | **Sequencing depth**  **[X]** | **N50**  **[bp]** |
| --- | --- | --- | --- |
| **Mix 5%** | 30.90 | 64035 | 16565.00 |
| **Mix 2%** | 30.80 | 66009 | 16565.00 |
| **Mix 1%** | 30.80 | 75179 | 16565.00 |
| **Mix 0.1%** | 30.90 | 70154 | 16565.00 |
| **Fibros_L-3086_P1** | 30.90 | 65663 | 16564.00 |
| **Fibros_L-15695_P1** | 30.80 | 73651 | 16566.00 |
| **Fibros_L-15935_P1** | 30.90 | 91115 | 16565.00 |
| **Fibros_L-20393_P1** | 30.90 | 83801 | 16562.00 |
| **Fibros_L-3086_P4** | 30.90 | 83099 | 16564.00 |
| **Fibros_L-15695_P4** | 30.80 | 81542 | 16566.00 |
| **Fibros_L-15935_P4** | 30.90 | 88923 | 16565.00 |
| **Fibros_L-20393_P4** | 30.90 | 92396 | 16562.00 |
| **Fibros_L-3086_P8** | 30.90 | 81847 | 16564.00 |
| **Fibros_L-15695_P8** | 30.90 | 87691 | 16566.00 |
| **Fibros_L-15935_P8** | 30.80 | 95769 | 16565.00 |
| **Fibros_L-20393_P8** | 30.90 | 79959 | 16562.00 |
| **Fibros_L-3086_P12** | 31.00 | 86260 | 16564.00 |
| **Fibros_L-15695_P12** | 30.90 | 72746 | 16566.00 |
| **Fibros_L-15935_P12** | 30.90 | 78948 | 16565.00 |
| **Fibros_L-20393_P12** | 30.90 | 86547 | 16562.00 |
| **Fibros_L-3085_P1** | 31.00 | 107279 | 16564.00 |
| **Fibros_L-3085_P4** | 31.00 | 97562 | 16564.00 |
| **Fibros_L-3085_P8** | 31.00 | 99115 | 16564.00 |
| **Fibros_L-3085_P12** | 30.90 | 87028 | 16564.00 |

| **5% Mixture** | | | | | |
| --- | --- | --- | --- | --- | --- |
| **Fraction subsampling** | **Sequencing depth**  **[X]** | **Detected minor variants/expected minor variants** | **Additional variant calls** | **Sensitivity** | **F_1_ score** |
| 1.0 | 64,039X | 18/18 | 0 | 1 | 1 |
| 0.5 | 32,272X | 18/18 | 0 | 1 | 1 |
| 0.25 | 16,177X | 18/18 | 0 | 1 | 1 |
| 0.1 | 6,422X | 18/18 | 0 | 1 | 1 |
| 0.05 | 3135X | 18/18 | 0 | 1 | 1 |
| 0.025 | 1,524X | 18/18 | 0 | 1 | 1 |
| 0.01 | 582X | 18/18 | 0 | 1 | 1 |
| 0.005 | 311X | 18/18 | 0 | 1 | 1 |
| **2% Mixture** | | | | | |
| 1.0 | 66,013X | 18/18 | 0 | 1 | 1 |
| 0.5 | 33,187X | 18/18 | 0 | 1 | 1 |
| 0.25 | 16,608X | 18/18 | 1 | 1 | 0.99 |
| 0.1 | 6,650X | 18/18 | 1 | 1 | 0.99 |
| 0.05 | 3,231X | 18/18 | 1 | 1 | 0.99 |
| 0.025 | 1,572X | 18/18 | 1 | 1 | 0.99 |
| 0.01 | 651X | 18/18 | 0 | 1 | 1 |
| **1% Mixture** | | | | | |
| 1.0 | 75,183X | 18/18 | 0 | 1 | 1 |
| 0.5 | 37,744X | 18/18 | 1 | 1 | 0.99 |
| 0.25 | 18,893X | 18/18 | 1 | 1 | 0.99 |
| 0.1 | 7,545X | 18/18 | 1 | 1 | 0.99 |
| 0.05 | 3,864X | 17/18 | 2 | 0.98 | 0.97 |
| 0.025 | 1,871X | 18/18 | 0 | 1 | 1 |
| 0.01 | 743X | 18/18 | 1 | 1 | 0.99 |
| **0.1% Mixture** | | | | | |
| 1.0 | 70,158X | 18/18 | 10 | 1 | 0.91 |
| 0.5 | 34,781X | 16/18 | 9 | 0.96 | 0.90 |
| 0.25 | 17,413X | 17/18 | 12 | 0.98 | 0.87 |
| 0.1 | 7,008X | 18/18 | 7 | 1 | 0.93 |
| 0.05 | 3,505X | 0/18 | 4 | 0.65 | 0.76 |

**Supplementary Table 5. Performance of mtDNA heteroplasmy detection across sequencing depths in predefined haplotype mixtures following subsampling of PacBio HiFi sequencing data.**

F_1_ score = 2 × (precision × sensitivity) ÷ (precision + sensitivity); Sensitivity = (Number of true positive calls) ÷ (Number of true positive calls + Number of false negative calls); Precision = (Number of true positive calls) ÷ (Number of true positive calls + Number of false positive calls)

| **Position** | **Region/ Locus** | **Ref** | **Alt** | **Genomic context +/- 10 bp** | **Variant MITOMAP frequency** | **Variant level** | **Variant level (*Mitorsaw)*** |
| --- | --- | --- | --- | --- | --- | --- | --- |
| **299** | D-loop | C | A | AAATTTCCAC**-C-**AAACCCCCCC | 0.010% | 0.001 | 0.001 |
| **311** | D-loop | C | T | AACCCCCCCT**-C-**CCCCGCTTCT | 0.037% | 0.001 | - |
| **1419** | RNR1 | G | A | AGGGTCGAAG**-G-**TGGATTTAGC | 0.000% | 0.003 | 0.006 |
| **8078** | MT-CO2 | G | A | CTCATGAGCT**-G-**TCCCCACATT | 0.045% | 0.002 | - |
| **10063** | MT-ND3 | A | G | AGAGTAATAA**-A-**CTTCGCCTTA | No variant ref frequency available on UCSC | 0.001 | - |
| **10196** | MT-ND3 | C | G | CTATATCCCC**-C-**GCCCGCGTCC | No variant ref frequency available on UCSC | 0.001 | - |
| **10197** | MT-ND3 | G | C | TATATCCCCC**-G-**CCCGCGTCCC | 0.000 | 0.001 | - |
| **12467** | MT-ND5 | T | C | GCATCCACCT**-T-**TATTATCAGT | 0.002% | 0.005 | - |
| **15249** | MT-CYB | A | G | TGAGGAGGCT**-A-**CTCAGTAGAC | 0.002% | 0.001 | - |
| **15302** | MT-CYB | C | A | CTTCATCTTG**-C-**CCTTCATTAT | No variant ref frequency available on UCSC | 0.002 | - |

**Supplementary Table 6: Summary of mitochondrial variants discordant with the Illumina reference identified from the 0.1% mixture and regional relevance. All coordinates correspond to GRCh38/hg38 mitochondrial reference sequence in UCSC. MITOMAP frequencies are reported from GenBank full-length sequences.**

**Supplementary Table 7: Summary of common variants (i.e., shared variants between major and minor mixture components). Example data of the 5% mixture are displayed.**

| **Full data set (at 64,039X sequencing depth)** | | | |
| --- | --- | --- | --- |
| **Position** | **Reference** | **Variant** | **Variant level** |
| 73 | A | G | 1 |
| 263 | A | G | 1 |
| 489 | T | C | 1 |
| 750 | A | G | 1 |
| 1438 | A | G | 1 |
| 2706 | A | G | 1 |
| 3010 | G | A | 1 |
| 4769 | A | G | 1 |
| 7028 | C | T | 1 |
| 8860 | A | G | 1 |
| 10398 | A | G | 1 |
| 11719 | G | A | 1 |
| 14766 | C | T | 1 |
| 15326 | A | G | 1 |
| **Subsampled data set (0.005 fraction of sequenced data at 311X sequencing depth)** | | | |
| 73 | A | G | 1 |
| 263 | A | G | 1 |
| 489 | T | C | 1 |
| 750 | A | G | 1 |
| 1438 | A | G | 1 |
| 2706 | A | G | 0.997 |
| 3010 | G | A | 1 |
| 4769 | A | G | 1 |
| 7028 | C | T | 0.997 |
| 8860 | A | G | 1 |
| 10398 | A | G | 1 |
| 11719 | G | A | 1 |
| 14766 | C | T | 1 |
| 15326 | A | G | 1 |

**Supplementary Figure**
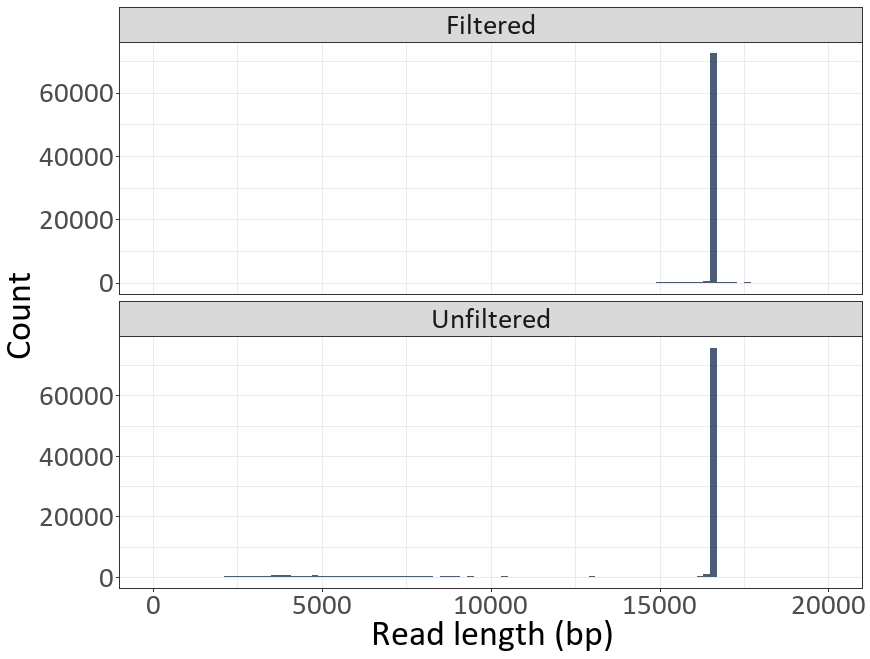
**1. Exemplary distribution of read lengths before and after filtering.** Histograms show the distribution of sequencing read lengths for unfiltered (bottom) and filtered (top) datasets. Read lengths are measured in base pairs (bp)

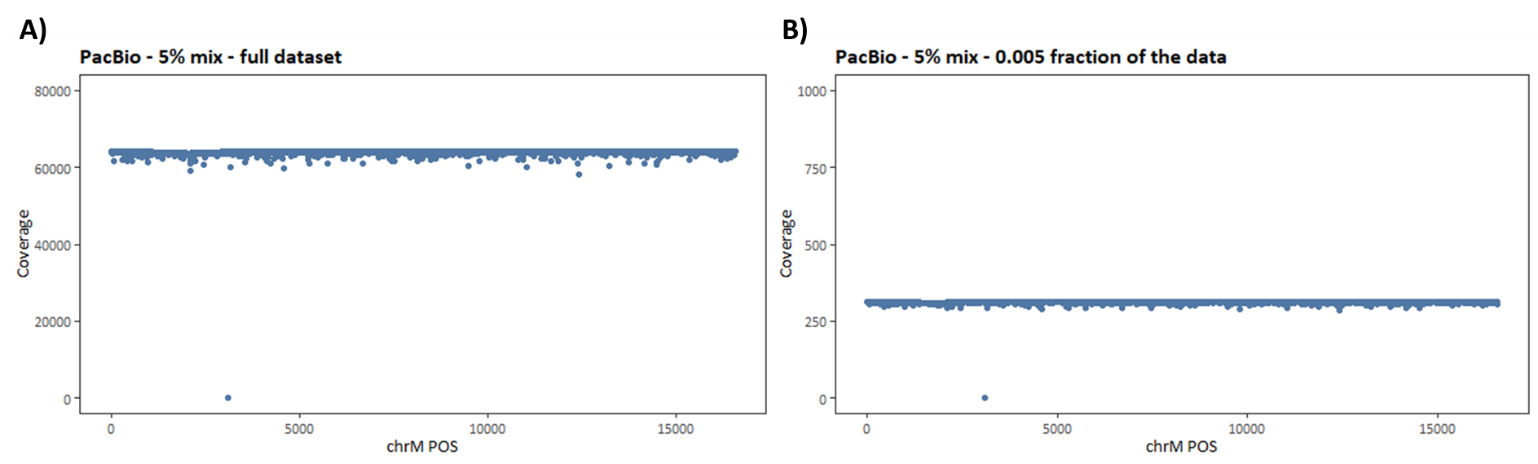

**Supplementary Figure 2. Sequencing depth across the mitochondrial genome, resulting in the single amplicon long-range PCR and PaBio sequencing.** Coverage across mitochondrial genome positions (chrM) for the 5% haplotype mixture using the full dataset (~64,000× depth, **(A)**) and after subsampling to ~300× depth **(B)**.

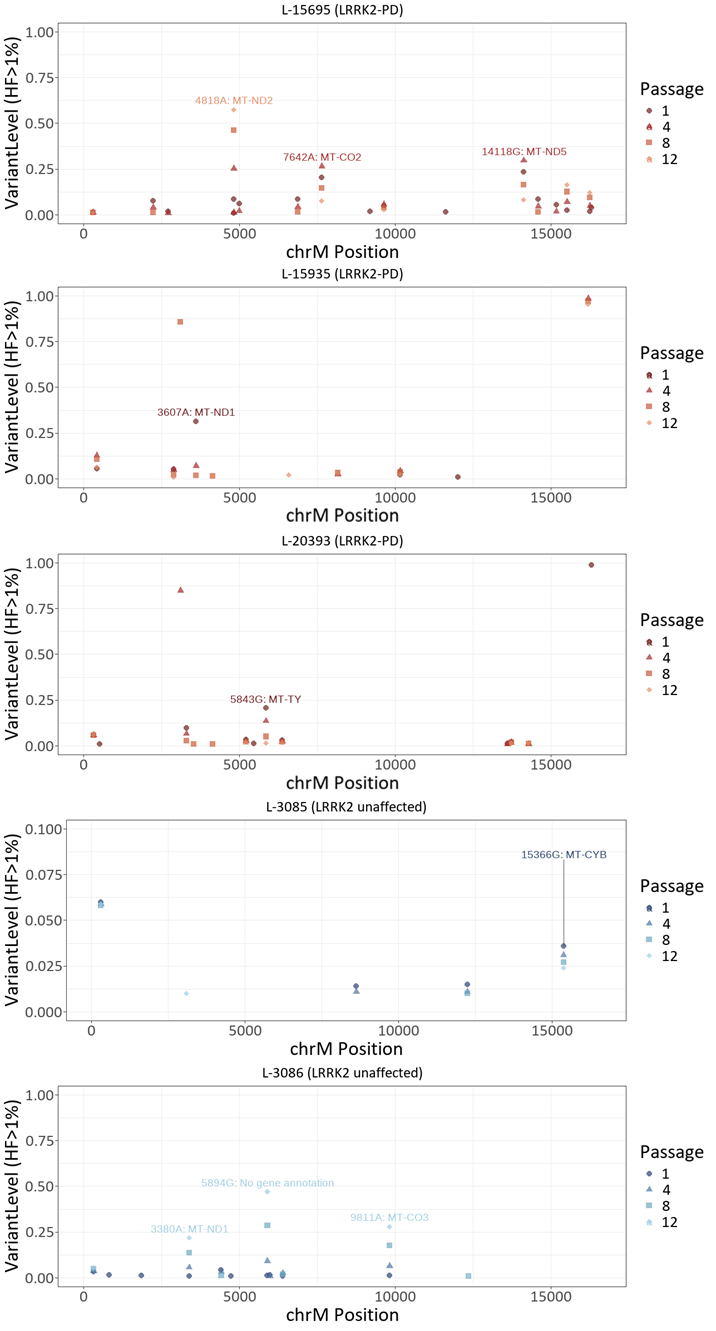
**Supplementary Figure 3.** **Heteroplasmy dynamics across fibroblast passages.** Heteroplasmic variants are plotted across mitochondrial genome positions for fibroblast cell lines derived from LRRK2 p.G2019S carriers. Panels represent individual cell lines, and symbols/colors indicate passages 1, 4, 8, and 12.
